# Trends in Mechanical Circulatory Support utilization, Left Ventricular Assist Device implantation and Transplant during Cardiogenic Shock Hospitalizations, after the New Heart Allocation Policy

**DOI:** 10.1101/2024.10.23.24316025

**Authors:** Diala Steitieh, Robert Beale, Ethan Katznelson, Elizabeth Feldman, Dilan Minutello, Daniel Lu, Parag Goyal, Jim Cheung, Luke K Kim, Udhay Krishnan

## Abstract

**Background:** In October 2018, a new heart allocation policy was implemented to risk stratify patients listed for transplant, prioritizing patients supported with temporary mechanical circulatory support (MCS). The policy changes may have had an impact on the management of cardiogenic shock (CS). We sought to determine the changes in use of temporary MCS, durable left ventricular assist device (LVAD) and transplant in patients hospitalized before and after the new policy.

**Methods:** A retrospective analysis was conducted using the National Inpatient Sample (NIS) between 2017– 2020. Hospitalizations for cardiogenic shock were identified, and stratified based on whether patients were admitted before or after the policy change. Baseline characteristics were compared between cohorts, and the primary outcome of interest was the use of MCS, transplant and LVAD before and after the policy change. Subgroup analyses included patients hospitalized at transplant and non-transplant centers, LVAD recipients as well as those who underwent transplant.

**Results:** A total of 643,655 hospitalizations were included, of which 260,340 (40.4%) were before the policy change, and 383,315 (59.6%) were after. In all patients with CS, there was a decrease in the use of LVAD (adjusted OR 0.73, p<0.01) and an increase in cardiac transplant (adjusted OR 1.45, p<0.01). While IABP use declined for the general CS population (adjusted OR 0.81, p<0.01), it increased significantly in cardiac transplant recipients (adjusted OR 2.55; p<0.01). Impella and VA-ECMO also increased in transplant recipients. No uptrend was seen in any other subgroup including LVAD recipients or CS patients managed in transplant centers.

**Conclusion:** Our study showed that the allocation policy change had a direct impact on MCS use in the first two years after implementation, but this effect was isolated to patients who underwent transplantation. It will be important to study how policy changes influence the management of other shock populations over time.

## Introduction

Despite advances in the therapeutic landscape, cardiogenic shock (CS) remains a challenging condition characterized by high rates of morbidity and mortality^1^. In recent years, many hospitals and healthcare systems have created multidisciplinary shock teams, often led by advanced heart failure (HF) and transplant cardiology specialists. These teams prioritize the early recognition of CS, interpret hemodynamic data, select which patients might benefit from early interventions, and utilize various treatment strategies including inotropes, diuretics and percutaneous mechanical circulatory support (MCS) devices, all with the ultimate goal of improving the chances of meaningful recovery and survival. However, given the complex and often refractory nature of this syndrome, one of the most important objectives is to identify patients who may not recover and who may be candidates for a durable left ventricular assist device (LVAD) or cardiac transplantation.

In October 2018, the United Network for Organ Sharing (UNOS) implemented a new heart transplant allocation policy with 6 tiers, prioritizing patients supported with temporary MCS over patients on inotropes alone or durable LVAD support^2^. While subsequent waitlist mortality and time to transplantation have clearly improved, there have also been unintended consequences. The use of LVAD as bridge to transplant (BTT) has declined and the number of transplants performed annually has increased^3,4^. Additionally, studies in this population have shown an abrupt increase in the use of intra-aortic balloon pump (IABP), with 28% of patients undergoing transplant with a IABP after the policy change compared to 8% prior to 2018^5^. This suggests that there has been a clear shift in both temporary and durable support strategies, at least for sicker patients awaiting transplant. However, in light of the increased role multidisciplinary shock teams now play in the management of CS, with a view toward preserving a clear path to transplant or LVAD if needed, it is possible that this shift may also impact a broader CS population. In addition, given the growing cross-collaboration between transplant and non-transplant centers through spoke and hub regional networks, potential shifts in mechanical support utilization may not necessarily be confined to transplant centers only.

Accordingly, we sought to determine the changes in use of temporary MCS, durable LVAD and transplant in patients hospitalized with cardiogenic shock before and after implementation of the new heart allocation policy, using a large nationwide database.

## Methods

A retrospective analysis was conducted using the National Inpatient Sample (NIS) between 2017 – 2020. The NIS represents roughly 8 million discharges per year and is maintained by the Agency for Health Care Quality and Research (AHRQ)^6^. Each patient record in the NIS contains information on the patient’s diagnoses and procedures performed during the hospitalization, based on International Classification of Diseases, Tenth Revision–Clinical Modification (ICD-10-CM) codes. Institutional Review Board approval and informed consent were not required for the study as all data collection was derived from a publicly available, de-identified administrative database. Discharges were weighted based on the sampling scheme to permit inferences for a nationally representative population^6^.

### Study Population

From January 2017 through December 2020, we identified hospitalizations for patients aged ≥ 18 years using ICD-10-CM codes for cardiogenic shock (Table S1). Hospitalizations where patients were admitted during October 2018 were excluded, as this was the month when the updated heart allocation policy was implemented. Hospitalizations with missing information on mortality, or in which the patient had an unknown admission month (Figure 1) were also excluded.

**Figure 1:**
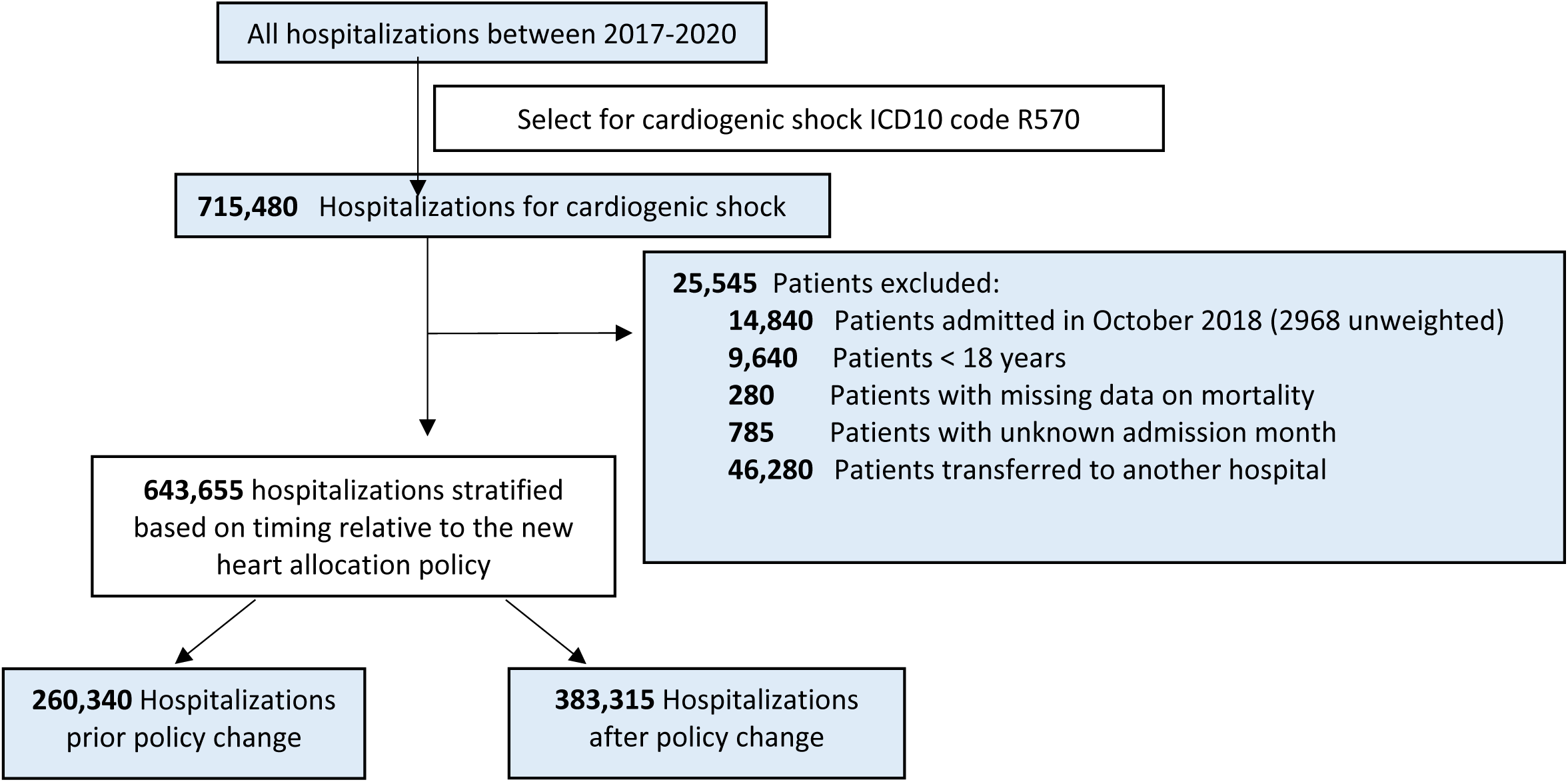
Study Cohort

### Primary Exposure and Outcome

Hospitalizations were stratified based on whether the patients were admitted before or after the 2018 policy change. The primary outcome of interest was the prevalence of temporary MCS, durable LVAD and heart transplant. Temporary MCS consisted of either extracorporeal membrane oxygenation (ECMO), Impella or IABP. These were identified using ICD-10-PR codes (Table S1). The use of Impella, IABP and ECMO were only captured if they occurred prior to LVAD placement or cardiac transplant, or anytime during the admission for patients who did not undergo LVAD placement or cardiac transplant.

### Statistical Analysis

Baseline characteristics were compared between patients admitted before and after the allocation policy change. Given the large sample size, standardized differences were calculated similar to previous studies of the NIS, with a difference >10% considered clinically meaningful. Continuous variables are presented as means; categorical variables are expressed as frequencies (percentages). For comparison of baseline characteristics, the Pearson chi-square tests was used for categorical variables and a survey-specific linear regression was used for continuous variables.

To determine whether a CS hospitalization after the allocation policy change was independently associated with use of temporary MCS, durable LVAD or transplant, we constructed a multivariable logistic regression model incorporating the following variables: age, sex, primary expected payer status, median household income, weekday versus weekend admission, hospital characteristics (region, bed size, location, and teaching status), all Elixhauser comorbidities, cardiac comorbidities (prior myocardial infarction, prior percutaneous coronary intervention (PCI), prior coronary artery bypass graft surgery (CABG), coronary artery disease, and family history of coronary artery disease), presentation (acute myocardial infarction (AMI) or non-AMI), and procedures (PCI, CABG). We repeated the analysis in certain subgroups including hospitalizations within transplant centers only, hospitalizations in which patient ultimately received LVADs or underwent transplant, hospitalizations in patients aged <=75 years, and patients with non-elective admissions. The effect of each subgroup on the relationship between IABP use and the NHAP was assessed by including interaction terms between the subgroups and IABP use.

For all analyses, we accounted for the complex survey design by using stratification and cluster variables. All analyses were conducted using STATA, version 15.1 (Statacorp, College Station, TX). All *P* values were 2 sided, with a significance threshold of *P*<0.05.

## Results

After applying the inclusion and exclusion criteria to the dataset, a total of 643,655 hospitalizations for cardiogenic shock were included in the final analysis. 260,340 hospitalizations (40.4%) occurred before the allocation policy change, and 383,315 (59.6%) were after the policy change (Figure 1). The baseline characteristics for the study population are displayed in Table 1. There were no significant differences in age or sex distribution between the cohorts (mean age 66.8 vs 66.7, p=0.82; 37.7% vs 37.8% women, p=0.71, respectively). There were similar rates of cardiovascular risk factors, including hypertension, prior MI, coronary artery disease, diabetes mellitus, obesity, chronic heart failure, valvular disease, and peripheral vascular disease (all standardized differences <10%). Rates of comorbidities were similar across all subgroups (see Supplemental tables). In the overall population, the rates of non–AMI-CS cases were significantly higher than AMI-CS cases (81.4% versus 18.6%), with a greater disparity after the allocation change (82.6% versus 17.4%; p <0.001; Table 2). Overall, revascularization (PCI or CABG) rates were low (less than 25% of the population).

**Table 1:** Baseline Characteristics for Patients Admitted with Cardiogenic Shock between 2017 and 2020, before and after the 2018 Allocation Policy Change.

| Characteristic | Overall <sup>a</sup> | Pre <sup>a</sup> | Post <sup>a</sup> | P Value <sup>b</sup> | Standardized Difference, % |
| --- | --- | --- | --- | --- | --- |
|  | 643,655 | 260,340 (40.4) | 383,315 (59.6) |  |  |
| Age, years | 66.7 | 66.8 | 66.7 | 0.82 <sup>c</sup> | 0.782 |
| Women | 242,830 (37.7) | 98,060 (37.7) | 144,770 (37.8) | 0.71 | 0.2151 |
| Black race | 101,820 (16.4) | 39,835 (15.9) | 61,985 (16.7) | <0.01 | <0.01 |
| Weekend admission | 151,980 (23.6) | 61,335 (23.6) | 90,645 (23.7) | 0.72 | 0.2074 |
| **Bed size** |  |  |  |  |  |
| Small | 82,895 (12.9) | 31,345 (12.0) | 51,550 (13.5) | <0.01 | 10.94 |
| Medium | 158,020 (24.6) | 64,450 (24.8) | 93,570 (24.4) |  | 1.41 |
| Large | 402,740 (62.6) | 164,545 (63.2) | 238,195 (62.1) |  | 1.7 |
| Urban location | 620,620 (96.4) | 250,965 (96.4) | 369,655 (96.4) | 0.72 | 0.2013 |
| Teaching hospital | 529,940 (82.3) | 210,035 (80.7) | 319,905 (83.5) | <0.01 | 7.2521 |
| **Region** |  |  |  |  |  |
| Northeast | 104,370 (16.2) | 41,530 (16.0) | 62,840 (16.4) | <0.01 | 2.72 |
| Midwest | 145,530 (22.6) | 59,360 (22.8) | 86,170 (22.5) |  | 1.41 |
| South | 257,340 (39.9) | 105,235 (40.4) | 152,105 (39.7) |  | 1.85 |
| West | 136,415 (27.2) | 54,215 (20.8) | 82,200 (21.4) |  | 2.92 |
| **Cardiac comorbidities** |  |  |  |  |  |
| Known CAD | 174,075 (27.0) | 72,430 (27.8) | 101,645 (26.5) | <0.01 | 2.9316 |
| Family history of CAD | 33,365 (5.2) | 14,450 (5.6) | 18,915 (4.9) | <0.01 | 2.7634 |
| Prior myocardial infarction | 75,065 (11.7) | 31,190 (12.0) | 43,875 (11.5) | <0.01 | 1.6615 |
| Prior percutaneous coronary intervention | 103,965 (16.2) | 45,240 (17.4) | 58,725 (15.3) | <0.01 | 5.5644 |
| Prior coronary artery bypass surgery | 56,510 (8.8) | 24,085 (9.3) | 32,425 (8.5) | <0.01 | 2.789 |
| **Elixhauser comorbidities** |  |  |  |  |  |
| Congestive heart failure | 411,670 (64.0) | 161,165 (62.0) | 250,505 (65.4) | <0.01 | 7.1692 |
| Chronic pulmonary disease | 84,515 (13.1) | 36,330 (14.0) | 48,185 (12.6) | <0.01 | 4.082 |
| Coagulopathy | 30,060 (4.7) | 11,835 (4.6) | 18,225 (4.8) | 0.08 | 0.9906 |
| Deficiency anemias | 40,590 (6.3) | 15,865 (6.1) | 24,725 (6.5) | 0.01 | 1.4698 |
| Diabetes mellitus (uncomplicated) | 52,600 (8.2) | 23,805 (9.1) | 28,795 (7.5) | <0.01 | 5.908 |
| Diabetes mellitus with complications | 206,610 (32.1) | 79,645 (30.6) | 126,965 (33.1) | <0.01 | 5.4325 |
| Hypertension | 86,980 (13.5) | 38,885 (14.9) | 48,095 (12.6) | <0.01 | 6.9434 |
| Liver disease | 154,205 (24.0) | 59,290 (22.8) | 94,915 (24.8) | <0.01 | 4.6706 |
| Fluid and electrolyte disorders | 440,420 (68.4) | 171,215 (65.8) | 269,205 (70.2) | <0.01 | 9.5822 |
| Obesity | 125,085 (19.4) | 48,455 (18.6) | 76,630 (20.0) | <0.01 | 3.4951 |
| Peripheral vascular disease | 37,765 (5.9) | 17,020 (6.5) | 20,745 (5.4) | <0.01 | 4.7503 |
| Pulmonary circulation disorder | 133,475 (20.7) | 52,705 (20.2) | 80,770 (21.1) | <0.01 | 2.0422 |
| Chronic renal failure | 249,710 (38.8) | 100,290 (38.5) | 149,420 (39.0) | 0.10 | 0.9407 |
| Valvular disease | 153,665 (23.9) | 62,830 (24.1) | 90,835 (23.7) | 0.07 | 1.0235 |
| Weight loss | 1,455 (0.2) | 580 (0.2) | 875 (0.2) | 0.84 | 0.1157 |

**a** Values are presented as number (percentage) of patients unless otherwise indicated.
**b** Rao-Scott $\chi^2$ test was used for all statistical tests in this table unless stated otherwise.
**c** Linear regression was performed.

**Table 2:** Case Presentations and the Use of MCS in All Patients With Cardiogenic Shock.

|  | **Overall <sup>a</sup>** | **Pre <sup>a</sup>** | **Post <sup>a</sup>** | **P value <sup>b</sup>** | **Standardized**<br>**Difference, %** |
| --- | --- | --- | --- | --- | --- |
| **Presentation** |  |  |  |  |  |
| AMI-CS | 119,670 (18.6) | 52,990 (20.4) | 66,680 (17.4) | <0.01 | 7.57 |
| Non-AMI CS | 523,985 (81.4) | 207,350<br>(79.7) | 316,635<br>(82.6) | <0.01 | 7.57 |
| Hospitalizations in<br>transplant centers | 160,440 (24.9) | 65,970 (25.3) | 94,470 (24.7) | <0.01 | 1.60 |
| **Interventions** |  |  |  |  |  |
| IABP | 78,110 (12.1) | 34,735 (13.3) | 43,375 (11.3) | <0.01 | 6.17 |
| Impella | 37,330 (5.8) | 14,760 (5.7) | 22,570 (5.9) | 0.10 | 0.94 |
| ECMO | 15,845 (2.5) | 6,350 (2.4) | 9,495 (2.5) | 0.67 | 0.25 |
| PCI or CABG | 155,680 (24.2) | 67,235 (25.8) | 88,445 (23.1) | <0.01 | 6.41 |
| LVAD | 4,890 (1.6) | 5,430 (1.9) | 10,320 (1.4) | <0.01 | 3.63 |
| Transplant | 2,050 (1.0) | 4,120 (0.8) | 6,170 (1.1) | <0.01 | 2.99 |

Differences in the use of use of temporary MCS, LVAD and transplant are illustrated in Figure 2. Across both periods, IABP was the predominant type of percutaneous circulatory support device. The prevalence of IABP use decreased slightly in the period after the policy change (13.3% of hospitalizations vs. 11.3%, p <0.01). Other percutaneous devices such as Impella and ECMO were used less frequently overall, with a similar rate before and after the allocation policy change (5.7% versus 5.9% for Impella, 2.4% versus 2.5% for ECMO). Cardiogenic shock hospitalizations in which cardiac transplantation was performed increased after the policy change, although this represented only 1% of the overall population. Approximately 25% of all cardiogenic shock hospitalizations took place in transplant centers. In these centers, the rates of cardiac transplantation increased from 3.1% to 4.4% (p < 0.01) after the policy change. Conversely, the frequency of LVAD decreased after the policy change in transplant centers (6.4% versus 4.6%, p < 0.01). After multivariable adjustment for the entire study population, IABP and LVAD use remained less prevalent in the period after the allocation policy change (adjusted OR 0.82 p<0.01, adjusted OR 0.73 p<0.01, respectively), and transplant remained more prevalent (adjusted OR 1.45, p<0.01). There was no significant difference in Impella and ECMO utilization.

**Figure 2:**
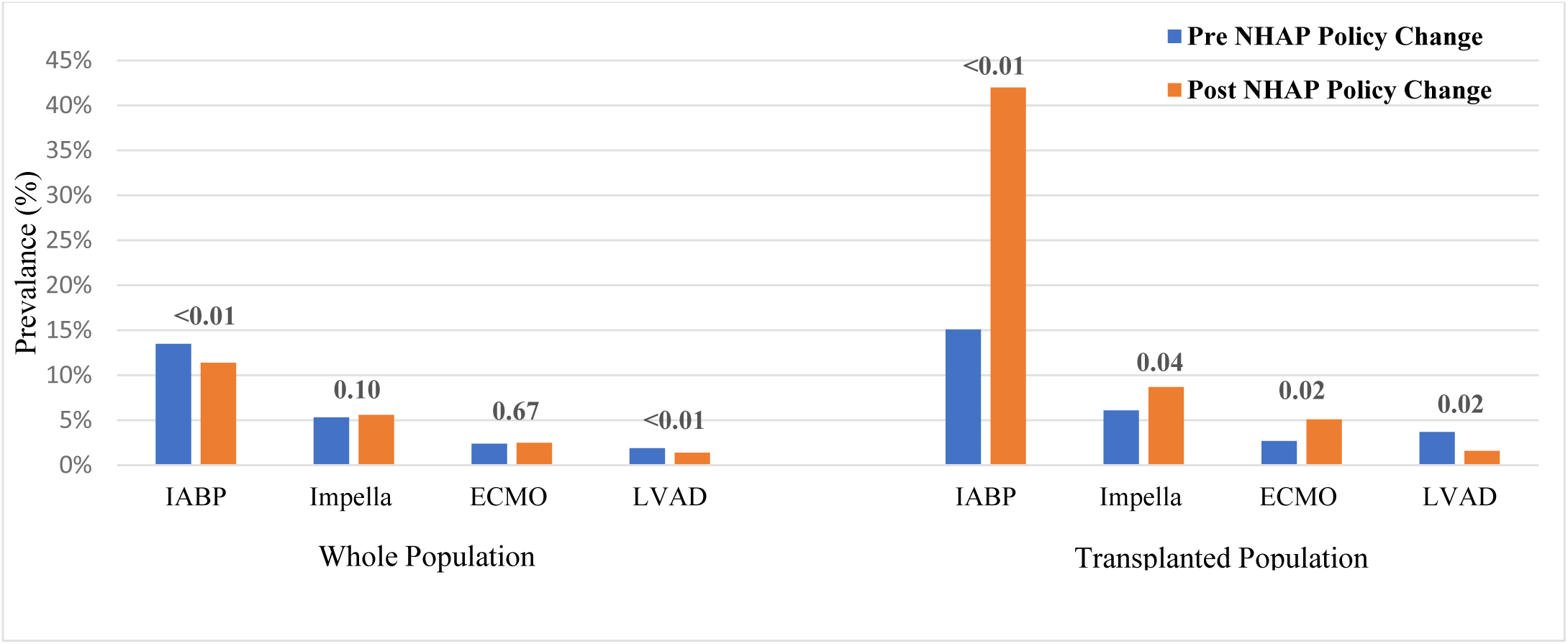
Frequency of MCS Use in Patients With Cardiogenic Shock, Pre and Post Policy Change

To better understand the impact of the policy change on temporary and durable therapy, we further evaluated additional subgroups. There were no changes in trends of device utilization in the subgroups stratified by acute coronary syndrome, nonelective admission status and age>75. However, there was a significant increase in the use of upstream IABP after the policy change in the cohort who underwent cardiac transplant during the hospitalization, which remained significant after multivariable adjustment (15.1% versus 41.6%, adjusted OR 2.55, p<0.01, Figure 3; p<0.01 for interaction). VA-ECMO and Impella prior to transplant also increased but were utilized far less frequently (2.7% versus 5.2%, adjusted OR 2.13 and 5.6% versus 8.7%, adjusted OR 1.73 respectively, p = 0.04 for both, Table 3). Lastly, in patients who received LVAD therapy, there was no difference in the use of ECMO, Impella or IABP after the policy change (see Supplemental Tables).

**Table 3:** Case Presentations and the Use of MCS in Patients Who Underwent Transplant.

|  | **Overall <sup>a</sup>** | **Pre <sup>a</sup>** | **Post <sup>a</sup>** | **P value <sup>b</sup>** | **Standardized Difference, %** |
| --- | --- | --- | --- | --- | --- |
| **Presentation** |  |  |  |  |  |
| ACS-CS | 85 (1.4) | 20 (1.0) | 65 (1.6) | 0.39 |  |
| Non-ACS CS | 6085 (98.6) | 2030 (99.0) | 4055 (98.4) | 0.39 | 5.36 |
| **Interventions** |  |  |  |  |  |
| IABP | 2025 (32.8) | 310 (15.1) | 1715 (41.6) | <0.01 | 61.46 |
| Impella/Tandem Heart | 475 (7.7) | 115 (5.6) | 360 (8.7) | 0.04 | 12.13 |
| ECMO | 270 (4.4) | 55 (2.7) | 215 (5.2) | 0.02 | 13.03 |
| LVAD | 140 (2.3) | 75 (3.7) | 65 (1.6) | 0.02 | 13.05 |

**Figure 3:**
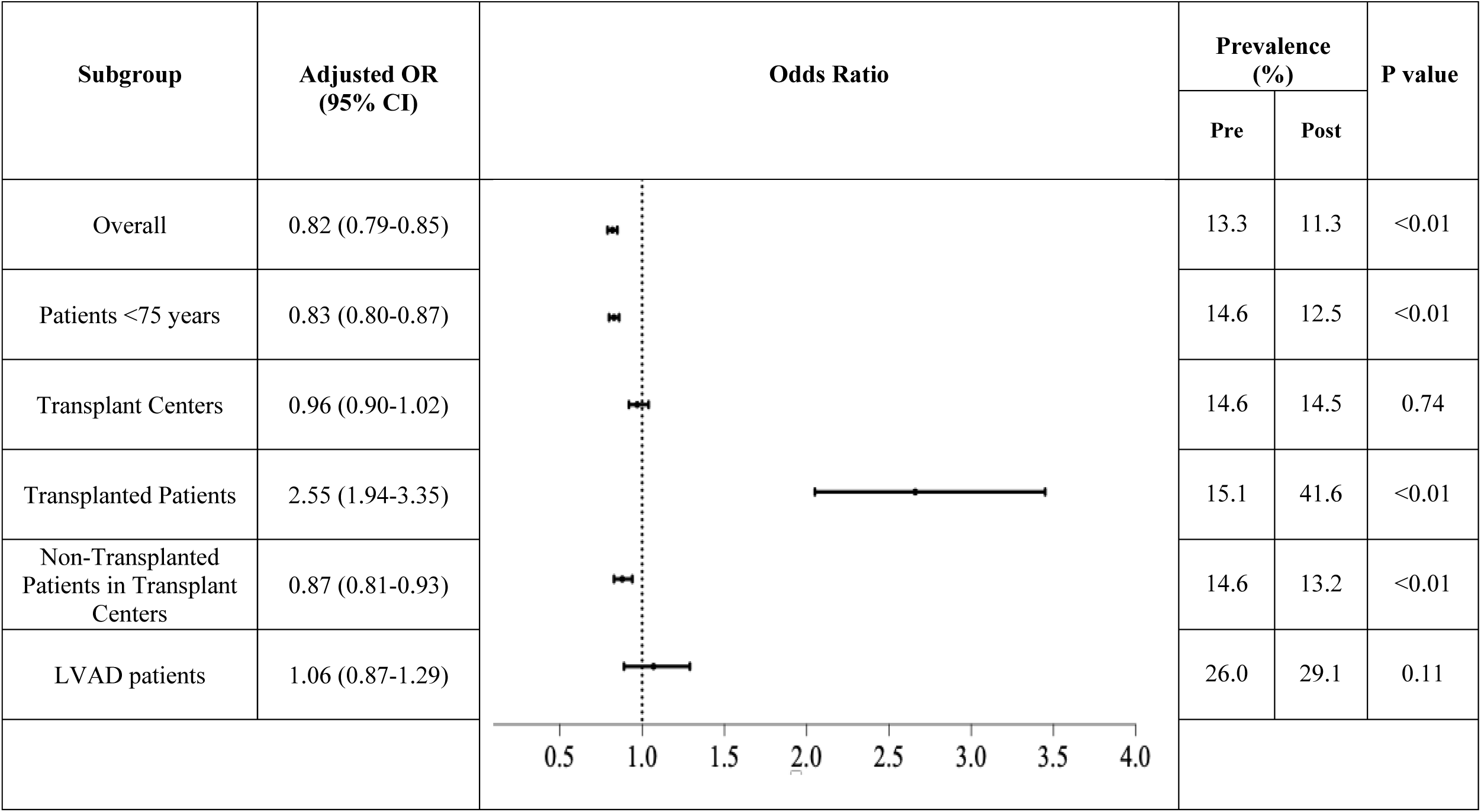
Odds of Receiving a IABP in Patients With Cardiogenic Shock After the policy change (With Hospitalizations Prior to the policy change as Reference)

## Discussion

While numerous studies have reported on changes in outcomes after the 2018 allocation policy implementation, most have focused only on listed patients that underwent transplant or LVAD. Given the advent of multidisciplinary shock teams led by HF physicians and vast hospital networks connected to transplant centers, we endeavored to look at the effect of the policy change on a broader CS population. In this large national study of CS hospitalizations, made up of mostly non-AMI-CS, roughly 25% took place in transplant centers. We found a decline in the use of LVADs and a rise in cardiac transplant after the policy change. While IABP use was relatively unchanged for the general CS population including LVAD recipients, it increased significantly in cardiac transplant recipients as did VA-ECMO and Impella, albeit to a lesser degree. These differences remained significant after multivariable adjustment for patient and hospital characteristics.

Our study found that allcomers with CS were less likely to undergo LVAD and more likely to undergo transplant, after the policy change. These findings are similar to the results of another recent study that found a 33% decrease in LVAD use in transplant candidates following the policy change^7^. The specific changes made in the 2018 listing criteria can help to elucidate these trends. Prior to October 2018, the allocation system was 3-tiered: 1A being highest priority, 1B intermediate priority and 2 the lowest priority. Previously, LVAD recipients were listed as status 1A for 30 days, at any point, once clinically stable after implantation and otherwise were listed as status 1B. The new allocation system expanded to a 6-tiered system to better risk stratify the heterogenous group of patients previously listed as 1A. In the new system, specific hemodynamic criteria within a 24-hour period were needed to meet at least status 2. In order to be considered status 1 or 2, patients with an LVAD would have to experience life threatening ventricular arrythmias or device malfunction/failure with the majority otherwise relegated to status 4^8^. Studies show that the most common listing status was 1B (74% of patients) before the policy change, and 2 (60% of patients) after the change^9^. As a consequence of all of this, the pathway from an LVAD implant to eventually receiving a transplant has become much less straightforward, which has dramatically affected patient and provider preferences regarding LVAD implantation. This is important, because it is clear that LVAD remains underutilized for patients with end stage heart failure^10^. In our study, LVAD use decreased to less than 5% of CS patients managed at transplant centers after the new allocation policy change. While some patients were, presumably, prioritized for transplant instead or didn’t have an indication for durable advanced therapies at the time, it’s possible that there were also end stage HF patients ineligible for transplant (for psychosocial reasons, comorbidities or increased pulmonary vascular resistance, etc.) who instead of undergoing LVAD, were managed medically in the hopes of preserving a pathway to transplant in the future. Because contemporary LVAD technology yields significant improvements in survival and functional capacity, such a delayed approach to advanced therapies might pose increased and unnecessary risks. The downstream effects of this de-prioritization of LVAD patients for transplant require further elucidation in subsequent studies.

The use of IABP for the management of CS has decreased significantly over the last twenty years, in part due to the lack of mortality benefit and increased short-term risks seen across multiple clinical trials^11,12,13^. This trend was recapitulated in our study. We found that the use of IABPs in all comers with CS continued to decline after the 2018 policy change. However, in CS admissions that did lead to cardiac transplantation, IABP utilization increased substantially after the change. Despite only 1% of the study population undergoing transplant, the rate of IABP escalation in this subgroup was enough to neutralize any decline in IABP use for CS, overall, at transplant centers. These findings are similar to a recent retrospective study of the UNOS database, which found that preoperative balloon pump use increased significantly following the policy change (7%–24.9%) in patients undergoing heart transplantation^14^. The higher IABP prevalence in our study (15.1% before and 41.6% after the policy change) likely reflects differences in the populations investigated, with our transplanted population being a hospitalized cohort enriched for patients that were likely status 2 or higher. It may have also included some CS patients treated and stabilized with IABP initially, who subsequently had it removed or were upgraded to another temporary MCS prior to transplant later in the hospitalization.

The observational nature of this study limits our ability to know with certainty why transplant recipients had higher upstream IABP use after the policy change but the reasons are likely multifactorial and nuanced. With the new allocation policy, a Status 2 listing was usually based on a need for temporary MCS such as IABP. Interestingly, a recent study showed that of 3,687 patients listed as Status 2 after the 2018 policy change, 48% were upgraded from a lower status indicating that many of these patients had chronic heart failure and were already listed at transplant centers prior to decompensating^15^. Therefore, many of the CS patients who underwent transplant in our study, were likely a unique group of shock patients, who had already partially adapted to a chronic low output state and were more likely to stabilize with IABP support alone^16^. Inta-aortic balloon pump use, specifically via an axillary approach, also allows for patients to be bridged for weeks or longer without compromising patient mobility and with a lower complication rate when compared to VA-ECMO or Impella^17^. These practical and physiologic benefits, which have been long recognized, but which the new allocation policy perhaps further illuminated, provides some context for the observed uptrend in IABP use. Finally, regarding outcomes, the implantation of an IABP has been shown to increase the likelihood of heart transplant^14^ without any signal for worsening post-transplant survival^18^. Despite all of this, it remains difficult to completely reconcile the rapid growth in IABP utilization for this one small shock cohort with the continued stagnation or decline in every other CS population over the same period. We agree with others that the observed disparity may also, at least in part, reflect a strategic effort in some cases to elevate candidate status and reduce waitlist time^19^.

Notably our study, which looked at a nationwide sample, did not show an uptrend in IABP use among the broader CS population, including all other subgroups managed in transplant centers as well as non-transplant centers. This is in contrast to a previous study of 7 transplant centers, which showed a significant increase in IABP use after the policy change, among non-AMI-CS patients suggesting a possible effect of the new policy on contemporary management in transplant centers^5^. Furthermore, while LVADs are still most commonly used as a rescue therapy for CS^20^, even in this enriched, high-acuity subgroup, there was no uptrend in IABP or other MCS use in the post allocation change era. Our study, therefore, showed that the allocation policy change has had a direct impact on IABP use, but that this effect has been isolated to patients who underwent transplantation, without clearly influencing the management of other populations. Still, as treatment paradigms continue to evolve over time with heart failure physicians increasingly managing shock in both hub and spoke centers, it will be important to continually study how the 2018 policy change may ultimately influence practice patterns for a broader shock population.

Impella and VA- ECMO were used less frequently overall in the CS population, with a similar rate before and after the policy change. As shown previously, ECMO use prior to transplant, did increase although to a much lesser degree compared to IABP^21^. Like IABP use after the policy change, bridging with ECMO to transplant has been shown to improve the frequency of heart transplant and decrease wait times^22^. Interestingly the overall use of ECMO (pre-operatively and post-operatively) in the transplant population did not statistically change because of less post-operative placement in the later era. While upstream placement would have eliminated the need to consider ECMO salvage post-operatively in some, the decreased rate of transplant for LVAD patients, was another likely explanation for this finding. In one study in the era prior to the policy change, 32% of patients with severe primary graft dysfunction requiring VA-ECMO had previously received an LVAD^23^. The modest decline in ECMO after heart transplant may represent one unintended, positive effect of the allocation change as salvage ECMO after heart transplant has correlated with increased mortality and resource utilization^24^. Impella use prior to transplant also increased only modestly after the policy change. Despite clear hemodynamic superiority compared to IABP^25^ and one recent study showing a mortality benefit in AMI-CS^26^, Impella use has correlated with a higher incidence of vascular complications, acute kidney injury, bleeding, hemolysis and stroke in CS trials^27^. Additionally, patients managed with an Impella are still assigned to the same Status 2 listing as those managed with IABP. The risk/benefit profile without affording any additional priority on the waitlist, as compared to IABP, may explain the relative underutilization of Impella in the post allocation change era. However, the Impella 5.5, surgically implanted via the axillary artery, can provide greater support for weeks or longer with a lower risk of complications compared to Impella CP or 2.5^28,29^. A recent UNOS analysis of the Impella 5.5 prior to transplant showed good waitlist and post-transplant outcomes with minimal device-related complications^30^. As comfort with this device continues to grow, future studies will likely confirm an expanded role for it in the management of patients listed for transplant as well as the broader CS population.

A major strength of this study was that we used a nationally representative cohort to capture the effect of a policy change on a real-world case mix of CS. However, there are notable limitations. First, while we were able to use ICD-10-CM codes to analyze data, miscoded and missing information can occur in large administrative datasets such as the NIS. However, HCUP quality control procedures are regularly performed to confirm that NIS data values are valid, consistent, and reliable^4^. Second, starting in 2012, the NIS was redesigned to represent a 20% national patient-level sample of all US hospitals. Because of the stratified sampling technique, it is possible that our definition of a transplant hospital may have missed some hospitals with a very low procedural volume (≤5 transplants annually). However, we have previously shown that advanced cardiac care hospitals, specifically hospitals that perform LVAD, can be identified in the NIS database with high accuracy^31^. Third, the NIS only includes data that can be encoded with ICD-10-CM codes, and therefore cannot include detailed information such as hemodynamic data, end organ function and degree of cardiomyopathy. Fourth, the NIS does not contain granular patient-level data, such as the use of medications, the baseline ejection fraction, laboratory values, and listing status for transplant. Fifth, the NIS provides short-term retrospective data, and therefore long-term outcomes following the NHAP cannot be assessed presently. Finally, because the post allocation era in this study also overlapped with the COVID 19 pandemic, this could have also affected MCS use, particularly in the year 2020. It will be important to continually follow trends in device use for CS, over time.

## Conclusion

In this large national study of CS hospitalizations, made up of mostly non-AMI-CS, there was a decrease in the use of LVADs and an increase in cardiac transplant after the 2018 allocation policy change. While IABP use declined for the general CS population, it increased significantly in cardiac transplant recipients. No uptrend was seen in any other subgroup including LVAD recipients or CS patients managed in transplant centers. While the impact of the policy change on management was isolated to the transplant population in the first 2 years after implementation, it will be important to continually study how it influences practice patterns over time for a broader shock population.

## Data Availability

Data is available from the National Inpatient Sample (NIS)

## Acknowledgements

None

## Sources of Funding

This work was supported by grants from the Michael Wolk Heart Foundation (New York, NY) and the New York Cardiac Center, Inc (New York, NY)). The Michael Wolk Heart Foundation and the New York Cardiac Center, Inc. had no role in the design and conduct of the study, in the collection, analysis, and interpretation of the data, or in the preparation, review, or approval of the manuscript.

## Disclosures

Dr Luke Kim has received fellowship grant support from Abbott and Medtronic.

Dr. Jim Cheung has received honoraria from Abbott and Medtronic and fellowship grant support from Abbott and Medtronic.

## Supplemental Materials

Supplemental Tables 1-21

## Notes

### Author Declarations

Institutional Review Board approval and informed consent were not required for the study as all data collection was derived from a publicly available, de-identified administrative database.

## References

1. Jentzer JC. Understanding Cardiogenic Shock Severity and Mortality Risk Assessment. Circ Heart Fail. 2020;13:e007568. doi: 10.1161/CIRCHEARTFAILURE.120.007568

2. Adult heart allocation. Organ Procurement and Transplantation Network, Health Resources & Services Administration. 2018.

3. Kilic A, Mathier MA, Hickey GW, Sultan I, Morell VO, Mulukutla SR, Keebler ME. Evolving Trends in Adult Heart Transplant With the 2018 Heart Allocation Policy Change. JAMA Cardiol. 2021;6:159–167. doi: 10.1001/jamacardio.2020.4909

4. Parker WF, Chung K, Anderson AS, Siegler M, Huang ES, Churpek MM. Practice Changes at U.S. Transplant Centers After the New Adult Heart Allocation Policy. J Am Coll Cardiol. 2020;75:2906–2916. doi: 10.1016/j.jacc.2020.01.066

5. Varshney AS, Berg DD, Katz JN, Baird-Zars VM, Bohula EA, Carnicelli AP, Chaudhry SP, Guo J, Lawler PR, Nativi-Nicolau J, et al. Use of Temporary Mechanical Circulatory Support for Management of Cardiogenic Shock Before and After the United Network for Organ Sharing Donor Heart Allocation System Changes. JAMA Cardiol. 2020;5:703–708. doi: 10.1001/jamacardio.2020.0692

6. Overview of the Healthcare Cost and Utilization Project (HCUP) National Inpatient Sample (NIS). In: Healthcare Cost and Utilization Project. Agency for Healthcare Research and Quality; 2023.

7. Mullan CW, Chouairi F, Sen S, Mori M, Clark KAA, Reinhardt SW, Miller PE, Fuery MA, Jacoby D, Maulion C, et al. Changes in Use of Left Ventricular Assist Devices as Bridge to Transplantation With New Heart Allocation Policy. JACC Heart Fail. 2021;9:420–429. doi: 10.1016/j.jchf.2021.01.010

8. Shore S, Golbus JR, Aaronson KD, Nallamothu BK. Changes in the United States Adult Heart Allocation Policy: Challenges and Opportunities. Circ Cardiovasc Qual Outcomes. 2020;13:e005795. doi: 10.1161/CIRCOUTCOMES.119.005795

9. Liu J, Yang BQ, Itoh A, Masood MF, Hartupee JC, Schilling JD. Impact of New UNOS Allocation Criteria on Heart Transplant Practices and Outcomes. Transplant Direct. 2021;7:e642. doi: 10.1097/TXD.0000000000001088

10. Tedford RJ, Leacche M, Lorts A, Drakos SG, Pagani FD, Cowger J. Durable Mechanical Circulatory Support: JACC Scientific Statement. J Am Coll Cardiol. 2023;82:1464–1481. doi: 10.1016/j.jacc.2023.07.019

11. Nishimoto Y, Inohara T, Kohsaka S, Sakakura K, Kawai T, Kikuchi A, Watanabe T, Yamada T, Fukunami M, Yamaji K, et al. Changing Trends in Mechanical Circulatory Support Use and Outcomes in Patients Undergoing Percutaneous Coronary Interventions for Acute Coronary Syndrome Complicated With Cardiogenic Shock: Insights From a Nationwide Registry in Japan. J Am Heart Assoc. 2023;12:e031838. doi: 10.1161/JAHA.123.031838

12. Thiele H, Zeymer U, Neumann FJ, Ferenc M, Olbrich HG, Hausleiter J, Richardt G, Hennersdorf M, Empen K, Fuernau G, et al. Intraaortic balloon support for myocardial infarction with cardiogenic shock. N Engl J Med. 2012;367:1287–1296. doi: 10.1056/NEJMoa1208410

13. Nan Tie E, Dinh D, Chan W, Clark DJ, Ajani AE, Brennan A, Dagan M, Cohen N, Oqueli E, Freeman M, et al. Trends in Intra-Aortic Balloon Pump Use in Cardiogenic Shock After the SHOCK-II Trial. Am J Cardiol. 2023;191:125–132. doi: 10.1016/j.amjcard.2022.12.019

14. Huckaby LV, Seese LM, Mathier MA, Hickey GW, Kilic A. Intra-Aortic Balloon Pump Bridging to Heart Transplantation: Impact of the 2018 Allocation Change. Circ Heart Fail. 2020;13:e006971. doi: 10.1161/CIRCHEARTFAILURE.120.006971

15. Pahwa S, Trivedi JR, Slaughter M. Status 2 Listing Strategies for Heart Transplantation: IABP or Impella? The Journal of Heart and Lung Transplantation. 2022;41:S387. doi: 10.1016/j.healun.2022.01.1534

16. Baldetti L, Pagnesi M, Gramegna M, Belletti A, Beneduce A, Pazzanese V, Calvo F, Sacchi S, Van Mieghem NM, den Uil CA, et al. Intra-Aortic Balloon Pumping in Acute Decompensated Heart Failure With Hypoperfusion: From Pathophysiology to Clinical Practice. Circ Heart Fail. 2021;14:e008527. doi: 10.1161/CIRCHEARTFAILURE.121.008527

17. Naqvi SY, Salama IG, Yoruk A, Chen L. Ambulatory Intra Aortic Balloon Pump in Advanced Heart Failure. Card Fail Rev. 2018;4:43–45. doi: 10.15420/cfr.2018:22:1

18. O’Connell G, Wang AS, Kurlansky P, Ning Y, Farr MA, Sayer G, Uriel N, Naka Y, Takeda K. Impact of UNOS allocation policy changes on utilization and outcomes of patients bridged to heart transplant with intra-aortic balloon pump. Clin Transplant. 2022;36:e14533. doi: 10.1111/ctr.14533

19. Trivedi JR, Slaughter MS. “Unintended” Consequences of Changes in Heart Transplant Allocation Policy: Impact on Practice Patterns. ASAIO J. 2020;66:125–127. doi: 10.1097/MAT.0000000000001128

20. Yuzefpolskaya M, Schroeder SE, Houston BA, Robinson MR, Gosev I, Reyentovich A, Koehl D, Cantor R, Jorde UP, Kirklin JK, et al. The Society of Thoracic Surgeons Intermacs 2022 Annual Report: Focus on the 2018 Heart Transplant Allocation System. Ann Thorac Surg. 2023;115:311–327. doi: 10.1016/j.athoracsur.2022.11.023

21. Golbus JR, Gupta K, Colvin M, Cascino TM, Aaronson KD, Kumbhani DJ, Saran R, Nallamothu BK. Changes in Type of Temporary Mechanical Support Device Use Under the New Heart Allocation Policy. Circulation. 2020;142:1602–1604. doi: 10.1161/CIRCULATIONAHA.120.048844

22. Gonzalez MH, Acharya D, Lee S, Leacche M, Boeve T, Manandhar-Shrestha N, Jovinge S, Loyaga-Rendon RY. Improved survival after heart transplantation in patients bridged with extracorporeal membrane oxygenation in the new allocation system. J Heart Lung Transplant. 2021;40:149–157. doi: 10.1016/j.healun.2020.11.004

23. Jacob S, Lima B, Gonzalez-Stawinski GV, El-Sayed Ahmed MM, Patel PC, Belli EV, Makey IA, Thomas M, Landolfo K, Landolfo C, et al. Extracorporeal membrane oxygenation as a salvage therapy for patients with severe primary graft dysfunction after heart transplant. Clin Transplant. 2019;33:e13538. doi: 10.1111/ctr.13538

24. Krishnamoorthy B, Mehta V, Critchley W, Callan P, Shaw S, Venkateswaran R. Financial implications of using extracorporeal membrane oxygenation following heart transplantation. Interact Cardiovasc Thorac Surg. 2021;32:625–631. doi: 10.1093/icvts/ivaa307

25. De Lazzari B, Capoccia M, Badagliacca R, Bozkurt S, De Lazzari C. IABP versus Impella Support in Cardiogenic Shock: “In Silico” Study. J Cardiovasc Dev Dis. 2023;10. doi: 10.3390/jcdd10040140

26. Moller JE, Engstrom T, Jensen LO, Eiskjaer H, Mangner N, Polzin A, Schulze PC, Skurk C, Nordbeck P, Clemmensen P, et al. Microaxial Flow Pump or Standard Care in Infarct-Related Cardiogenic Shock. N Engl J Med. 2024;390:1382–1393. doi: 10.1056/NEJMoa2312572

27. Thakkar S, Patel HP, Kumar A, Tan BE, Arora S, Patel S, Doshi R, Depta JP, Kalra A, Dani SS, et al. Outcomes of Impella compared with intra-aortic balloon pump in ST-elevation myocardial infarction complicated by cardiogenic shock. Am Heart J Plus. 2021;12:100067. doi: 10.1016/j.ahjo.2021.100067

28. Bhardwaj A, Salas I, Al Rameni D, Patel M, Akay M, Kar B, Gregoric I. Utilization of impella 5.5 in patients with cardiogenic shock as a bridge to decision, recovery, or destination therapy. IHJ Cardiovascular Case Reports (CVCR). 2023;7. doi: 10.1016/j.ihjccr.2023.06.002

29. Lewin D, Nersesian G, Lanmueller P, Falk V, Potapov EV, Ott S. Long-Term Complications After Support with Impella 5.0 and 5.5. The Journal of Heart and Lung Transplantation. 2022;41:S273–S274. doi: 10.1016/j.healun.2022.01.672

30. Cevasco M, Shin M, Cohen W, Helmers MR, Weingarten N, Rekhtman D, Wald JW, Iyengar A. Impella 5.5 as a bridge to heart transplantation: Waitlist outcomes in the United States. Clin Transplant. 2023;37:e15066. doi: 10.1111/ctr.15066

31. Wang JI, Lu DY, Mhs, Feldman DN, McCullough SA, Goyal P, Karas MG, Sobol I, Horn EM, Kim LK, et al. Outcomes of Hospitalizations for Cardiogenic Shock at Left Ventricular Assist Device Versus Non-Left Ventricular Assist Device Centers. J Am Heart Assoc. 2020;9:e017326. doi: 10.1161/JAHA.120.017326

